# How does palliative care fit into national health spending? A secondary analysis of trends in long-term healthcare expenditure in the United Kingdom

**DOI:** 10.64898/2026.01.23.26344608

**Authors:** Elisha De-Alker, Annette Alcock, Fliss E.M. Murtagh

## Abstract

**Objectives:** Current methods of health expenditure reporting make spending on palliative care services difficult to quantify. This paper (1) examines trends in the components of government (public) spending on health-related long-term care reported in the UK Health Accounts for the period of 2013 to 2022 to establish the wider context of palliative care expenditure, and (2) relates these trends to existing knowledge of expenditure on specialist palliative care services in the UK.

**Methods:** We conducted a descriptive secondary analysis of annually reported government expenditure on health-related long-term care between 2013 and 2022 from the UK Health Accounts dataset. We contrasted this with UK governmental and non-governmental spending on specialist palliative care services using annual expenditure figures reported by Hospice UK.

**Results:** Real-terms UK government spending on health-related long-term care grew by £6.4 billion (22.9%) between 2013 and 2022, from £27.9 to £34.3 billion. Real-terms spending on specialist palliative care grew by £110 million (10.7%) over the same period, from £1,027 to £1,137 million.

In 2022, spending on inpatient care comprised the majority of government health-related long-term care expenditure (£22.6 billion; 65.9%). Home-based care comprised one-third (£11.8 billion; 33.4%). Outpatient care accounted for 0.7% (£260.2 million). Equivalent data was not available for analysis of specialist palliative care expenditure.

**Conclusions:** Low granularity of UK national health expenditure accounts limits national and international comparisons of spending on palliative care. However, it is clear that UK expenditure on specialist palliative care services has not kept pace with growth in expenditure on health-related long-term care.

**What is already known on this topic:**

- Global demand for palliative care is increasing as rates of serious life-limiting illness, dementia, cancer and multiple long-term conditions rise internationally.
- Increasing complexity of illness and population ageing are two factors implicated in both rising healthcare expenditure and growing demand for palliative care internationally.
- The UK has previously been ranked as providing the highest quality of palliative care amongst international competitors – however, concerns about the longevity of funding sources for specialist palliative care services has led to calls for further investment.

**What this study adds:**

- Real terms UK government spending on health-related long-term care – which includes, but is not limited to, palliative care services – increased by 22.9% between 2013 and 2022.
- Over the same period, UK spending on specialist palliative care services as reported by Hospice UK grew by only 10.7%.
- Our results take into account health-related social care spending, which forms a key part of care for people living with illness, including those receiving palliative care services.

**How this study might affect research, practice or policy:**

- The future of funding for specialist palliative care in the UK is uncertain, and current funding frameworks are complex. This paper adds to ongoing policy discussions surrounding this issue, highlighting the discrepancy between growth in public sector spending on health-related long-term care and overall spending on specialist palliative care services (from governmental and non-governmental sources).

## INTRODUCTION

As the global burden of serious life-limiting diseases increases, an understanding of current expenditure on palliative care services is required in order to plan for the necessary expansion of health and social care services. [1, 2] Growth in healthcare expenditure as a proportion of world income poses a challenge for healthcare systems internationally. In the United Kingdom (UK), increases in healthcare expenditure have outpaced growth in Gross Domestic Product (GDP; see *Box 1*) over the last decade, a trend also observed across most other European countries. [3] UK healthcare expenditure has more than doubled as a share of national resources since the establishment of its National Health Service (NHS) in 1948. [4] This growth is influenced by a combination of demographic and non-demographic factors. Key drivers of healthcare expenditure include changing population age distributions, technological advancements in healthcare, and policy decisions. [5]

Demographic changes in the UK mirror those experienced by other high-income countries. The proportion of the UK population aged over 65 increased from 16.4% in 2011 to 18.6% in 2021, and the population older than 85 is expected to almost double from 1.6 million in 2018 to 3.0 million by 2043. [6, 7] These changes are accompanied by a marked expansion of multiple long-term conditions, associated with higher levels of complex care needs and healthcare costs. [8]

Included among this rising global burden of disease are serious life-threatening and life-limiting illnesses requiring palliative care services. An estimated 48 million people will die with serious health-related suffering by 2060, accounting for 47% of deaths globally. [1] The funding of palliative care services is therefore an economic priority. A 2015 comparison of 80 international healthcare systems ranked the UK as providing the best quality of palliative care, in part due to its relatively high levels of healthcare expenditure. [9] Despite this, concerns about the limitations of palliative care funding have led to calls for further investment. [10, 11]

It is estimated that up to 90% people who die each year in the UK could benefit from palliative care, and demand for these services is expected to rise with the increasing prevalence of cancer, dementia, and multiple long-term conditions. [12] Historically, national spending on palliative care has been difficult to quantify due to limited data availability. Current literature focuses on costs of care in the last year of life, rather than expenditure on palliative care provision as a whole. [11, 13]

Planning the equitable and effective distribution of palliative care requires an understanding of current expenditure on palliative care services. Palliative care is delivered by both specialists in palliative care and ‘non-specialists’ in palliative care (see *Box 1*). Specialist palliative care services, delivered by teams of multiprofessionals working across primary care, secondary care and hospice settings. They accept referrals from non-specialists and assess and treat the needs of people with life-limiting and life-limiting illnesses. Spending on specialist palliative care services should be interpreted in the wider context of health, social and informal care.

In the UK, annual healthcare expenditure is reported to internationally standardised definitions in the UK Health Accounts. [14] This methodology situates spending on palliative care within the category of health-related long-term expenditure. [15] Using the UK as an example, the objectives of this paper are (1) to examine trends in the components of government (public) spending on health-related long-term care reported in the UK Health Accounts for the period of 2013 to 2022 to establish the wider context of palliative care expenditure, and (2) to relate these trends to existing knowledge of expenditure on specialist palliative care services in the UK.

## METHODS

### Design

This study had two components: (1) a descriptive secondary analysis of annual government expenditure on nationally reported health-related long-term care for the period of 2013 to 2022 in the UK, and (2) comparison with identified government expenditure on health-related long-term care to government (and non-government) financed expenditure on specialist palliative care services.

### Data sources

1. *UK Health Accounts dataset: 2023 edition*. [16] https://www.ons.gov.uk/peoplepopulationandcommunity/healthandsocialcare/healthcaresystem/datasets/healthaccountsreferencetables
2. *Annual hospice charitable expenditure data provided by Hospice UK*. Also available in the annual Hospice UK Accounts, reported each year. [17] https://www.hospiceuk.org/innovation-hub/support-for-your-role/non-clinical-resources/finance-support-for-hospices/financial-benchmarking#content-menu-7843)

### Data definitions and inclusion

#### (1) UK Health Accounts

The UK’s nationally reported health expenditure data is presented in the UK Health Accounts. This dataset is produced to internationally standardised System of Health Accounts 2011 definitions, allowing comparison of expenditure between different national healthcare systems. The 2023 dataset was chosen as it was the most recently published complete dataset available. The System of Health Accounts, and therefore the UK Health Accounts, report health expenditure by healthcare function, provider and financing scheme. A summary of inclusion decisions is presented in. Further details of UK Health Accounts methodology can be found in *Appendix 1 (supplemental material)*.

##### Healthcare function

Palliative care expenditure is reported within the healthcare function ‘health-related long-term care’. Health-related long-term care includes medical and nursing services directed towards symptom management, administering medication, performing medical diagnoses and minor surgery, dressing wounds, health counselling, and providing emotional and spiritual support. Health-related long-term care therefore encompasses both specialist and non-specialist palliative care but extends well beyond it. It should be noted that health-related long-term care includes elements of personal care services conventionally considered social care, where support with basic Activities of Daily Living is required due to an underlying health condition. [18] Definitions of terms used in this paper are presented in *Table 1*.

**Table 1:** Definitions of key terms used in this paper.

| **Term** | **Definition** |
| --- | --- |
| Gross domestic product | A measure of a country’s economic growth based on the value of goods and services produced within a country over a given period. [19] |
| Health-related long-term care | Medical and personal care services consumed with the primary goal of alleviating pain and suffering, and reducing or managing the deterioration in health status in patients with a degree of long-term dependency. Examples of included services are age- and disability-related conditions, and palliative care. [14, 20] |
| Specialist palliative care | Healthcare services aimed at providing treatment and support for individuals with advanced life-limiting illnesses delivered by |
|  | healthcare professionals who predominantly practice in this field in community/primary care, hospice and hospital/secondary care settings. [21] |
| Non-specialist palliative care | Healthcare services aimed at providing treatment and support for individuals with advanced life-limiting illnesses delivered by medical professionals predominantly practicing in other specialist medical fields in both community/primary care and hospital/secondary care settings, for example general practitioners. [21] |

Other healthcare functions reported in the UK Health Accounts are curative care, rehabilitative care, ancillary services, (including ambulance and patient transport), medical goods (including pharmaceuticals, durable and non-durable goods, and therapeutic appliances), preventive care, and governance, health system financing and administration. These were excluded from our analysis as they did not include expenditure on palliative care.

##### Financing scheme

The UK Health Accounts categorises healthcare expenditure by funding source.

These include government, non-profit institutions serving households (charities), voluntary insurance schemes (voluntary insurance claims from private medical insurance and employer self-insurance schemes), enterprise (provision or purchase of healthcare services by companies for employees) and out-of-pocket payments (direct consumer spending on healthcare goods and services, such as prescription charges and direct payments for private healthcare services).

To meet our objective we selected government-financed expenditure for inclusion in our analysis as this included National Health Service and local authority spending on health-related long-term care, which included, but was not limited to, palliative care.

It should be noted that this government-financed expenditure does not include grants to charities such as Hospice UK.

Non-profit institutions play an important role in providing specialist palliative care in the UK hospice sector. However, UK Health Accounts methodological guidance highlights that reported charitable expenditure is taken from a representative and undisclosed sample of charities. Consequently, we could not conclude whether any charitable organisations providing palliative care services were included in this sample. We therefore excluded this category from our analysis.

##### Healthcare provider

We identified providers of health-related long-term care who deliver services including, but not solely limited to, palliative care for inclusion in our descriptive analysis. Selected providers included hospitals and long-term care facilities providing ‘inpatient’ care, general practices (GPs) and hospitals providing ‘outpatient’ care, and home healthcare agencies and community healthcare teams providing ‘home-based care’.

We excluded providers whose services did not directly include palliative care. These providers generally had no reported long-term healthcare expenditure (dental practice, ambulance services, retailers of medical goods, providers of preventive care, administration and financing). We also excluded households as providers of home healthcare, which covered only households in receipt of Carer’s Allowance, as expenditure on palliative care within this group could not be clearly defined. We excluded hospital day services, as secondary palliative care is generally delivered in inpatient or clinic (outpatient) services rather than as day case activity in hospitals.

In order to identify trends in annual health-related long-term care expenditure over time, real terms figures adjusted to 2023 prices were used, as reported in the 2023 edition of the UK Health Accounts dataset, to account for the effect of inflation.

#### (2) Annual hospice charitable expenditure via Hospice UK

Data provided by Hospice UK reflects spending on charitable purposes (direct patient care and education) by 190 member organisations. This includes spending by hospice charities on inpatient, outpatient, and community/home-based specialist palliative care services.

This expenditure incorporates both governmental and non-governmental expenditure, financed by charitable income and government grants to hospice charities.

Of note, this data does not include expenditure by hospital advisory specialist palliative care; in the UK, there are hospital advisory specialist palliative care teams in most acute hospitals.

### Data analysis

1. We performed descriptive analyses of UK Health Accounts data on governmental health-related long-term care expenditure using real terms figures adjusted to 2023 prices using the GDP deflator (released May 2024). We sorted and analysed data using Microsoft Excel. We generated pivot tables, line graphs and treemaps to illustrate change in expenditure over time, and composition of health-related long-term care.
2. We identified specialist palliative care expenditure as reported by Hospice UK Accounts over the period of 2013 to 2022. Hospice UK figures were adjusted to 2023 prices using the Consumer Price Index (released February 2025). [22]

Using real terms figures removes the impact of inflation, supporting meaningful comparison of expenditure over time.

## RESULTS

### Contextual findings

The UK Health Accounts report that total national healthcare expenditure increased by £64.9 billion between 2013 and 2022, from £237.3 billion to £296.6 billion (*Figure 1A*). This includes both publicly and privately funded expenditure by the government, charitable sector, consumer, enterprise and insurance schemes across all healthcare functions included in the UK Health Accounts.

**Figure 1:**
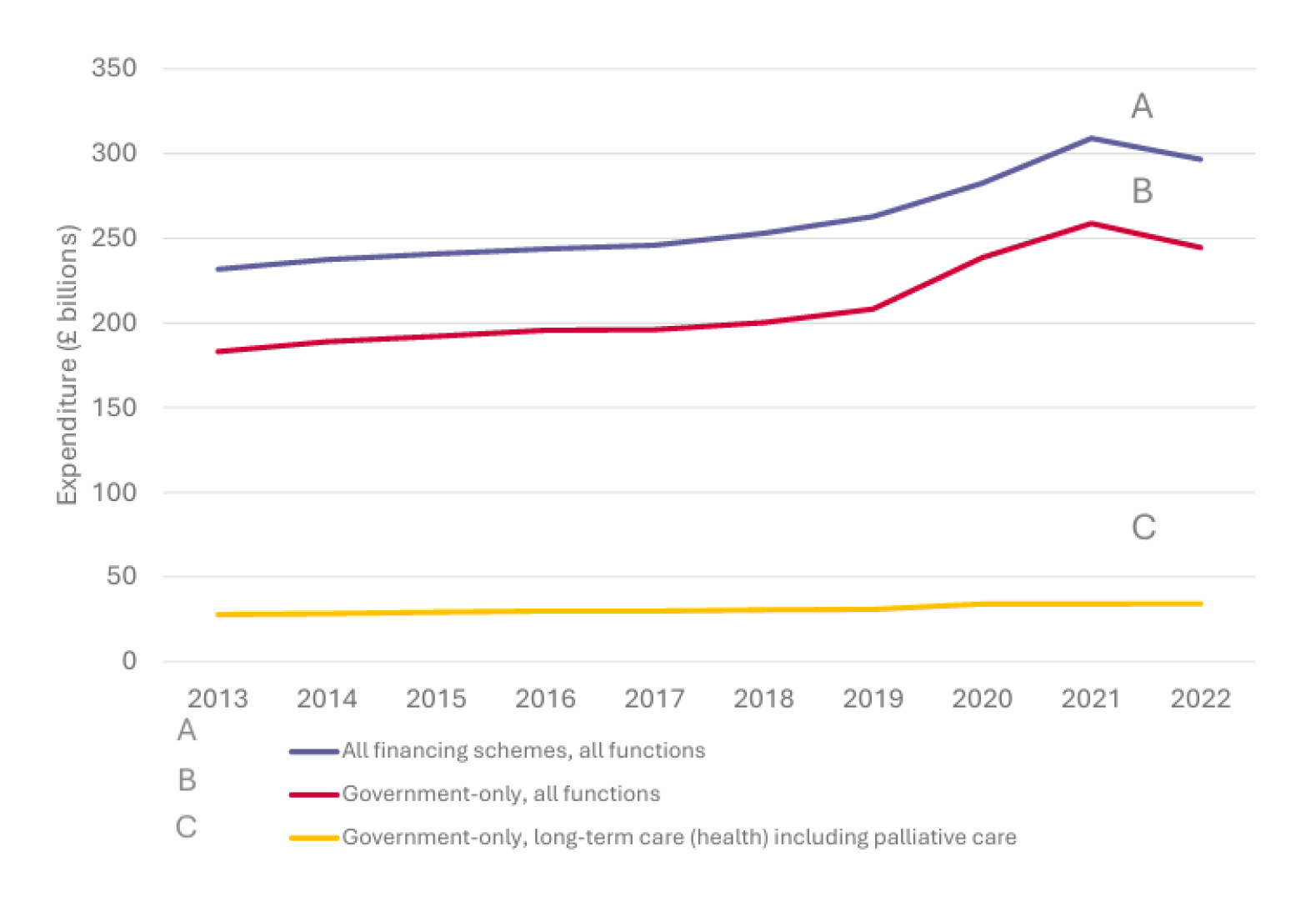
Graph representing annual UK healthcare expenditure from 2013 to 2022, adjusted for inflation to 2023 prices. (A) Annually reported total national healthcare expenditure by all financing schemes across all healthcare functions. (B) Annual government expenditure across all healthcare functions. (C) Annual government expenditure on health-related long-term care, delivered by selected providers (inpatient care provided by hospitals and long-term care facilities, outpatient care provided by GP surgeries and hospitals, and home-based care delivered by home care agencies and community nursing teams). This includes, but is not limited to, spending on palliative care. Source: UK Health Accounts.

Government (public) spending across all healthcare functions comprised, on average, 80.8% of this total national healthcare expenditure annually, and increased from £183.1 billion in 2013 to £244.5 billion in 2022 (*Figure 1B*). This includes spending by the NHS, local authorities and other government bodies on preventative, curative, rehabilitative and health-related long-term care, as well as pharmaceutical goods, medical devices, patient transport services and health system administration.

Both total national healthcare expenditure and government-financed healthcare expenditure increased year-on-year, accelerating due to the rollout of government-funded COVID-19 measures from 2020 until 2022, when spending decreased by £12.4 billion and £14.2 billion respectively with the reduction in vaccination and preventive schemes related to the COVID-19 pandemic. Detailed annual expenditure figures for total UK health expenditure, total government-financed health expenditure and government-financed health-related long-term care expenditure figures are presented in *Table 2*.

**Table 2:** Annual UK health expenditure for the period of 2013 to 2022 in real terms, adjusted to 2023 prices. Source: UK Health Accounts.

| Year | Total UK health expenditure<br>(across all financing schemes, healthcare functions and providers) | Total government-financed health expenditure<br>(across all financing schemes and providers) |  | Government-financed health-related long-term care expenditure<br>(selected providers) |  |  |
| --- | --- | --- | --- | --- | --- | --- |
|  | £ billions | £ billions | % of total UK health expenditure | £ billions | % of total UK health expenditure | % of government-financed health expenditure |
| 2013 | 231.7 | 183.1 | 79.0 | 27.9 | 12.0 | 15.2 |
| 2014 | 237.4 | 188.9 | 79.6 | 28.4 | 12.0 | 15.1 |
| 2015 | 240.9 | 192.1 | 79.8 | 29.3 | 12.2 | 15.2 |
| 2016 | 243.7 | 195.7 | 80.3 | 29.9 | 12.3 | 15.3 |
| 2017 | 245.8 | 196.0 | 79.8 | 29.9 | 12.2 | 15.3 |
| 2018 | 252.9 | 200.2 | 79.2 | 30.7 | 12.1 | 15.3 |
| 2019 | 262.8 | 208.2 | 79.2 | 30.9 | 11.8 | 14.8 |
| 2020 | 282.6 | 238.6 | 84.4 | 34.1 | 12.1 | 14.3 |
| 2021 | 309.0 | 258.7 | 83.7 | 34.2 | 11.1 | 13.2 |
| 2022 | 296.6 | 244.5 | 82.4 | 34.3 | 11.6 | 14.0 |

#### (1) Trends in national government-financed health-related long-term care expenditure and composition of health-related long-term care spend

Government-financed health-related long-term care expenditure increased from £27.9 billion in 2013 to £34.3 billion in 2022 (*Figure 1C*). This growth of £6.4 billion represents a proportional increase of 22.9% in government-financed health-related long-term care spend over the ten-year period analysed. Overall, spending on health-related long-term care increased year-on-year, comprising, on average, 11.9% of total national healthcare expenditure and 14.8% of total government-financed healthcare expenditure annually.

*Figure 2* illustrates the composition of total national healthcare expenditure, using 2022 as a reference year. Of the £296.6 billion spent on healthcare by all financing schemes across all healthcare functions in the UK in 2022, total government expenditure on healthcare accounted for £244.5 billion (*Figure 2B)*. Government expenditure on health-related long-term care comprised £34.3 billion of this total government spending on healthcare *(Figure 2C)*.

**Figure 2:**
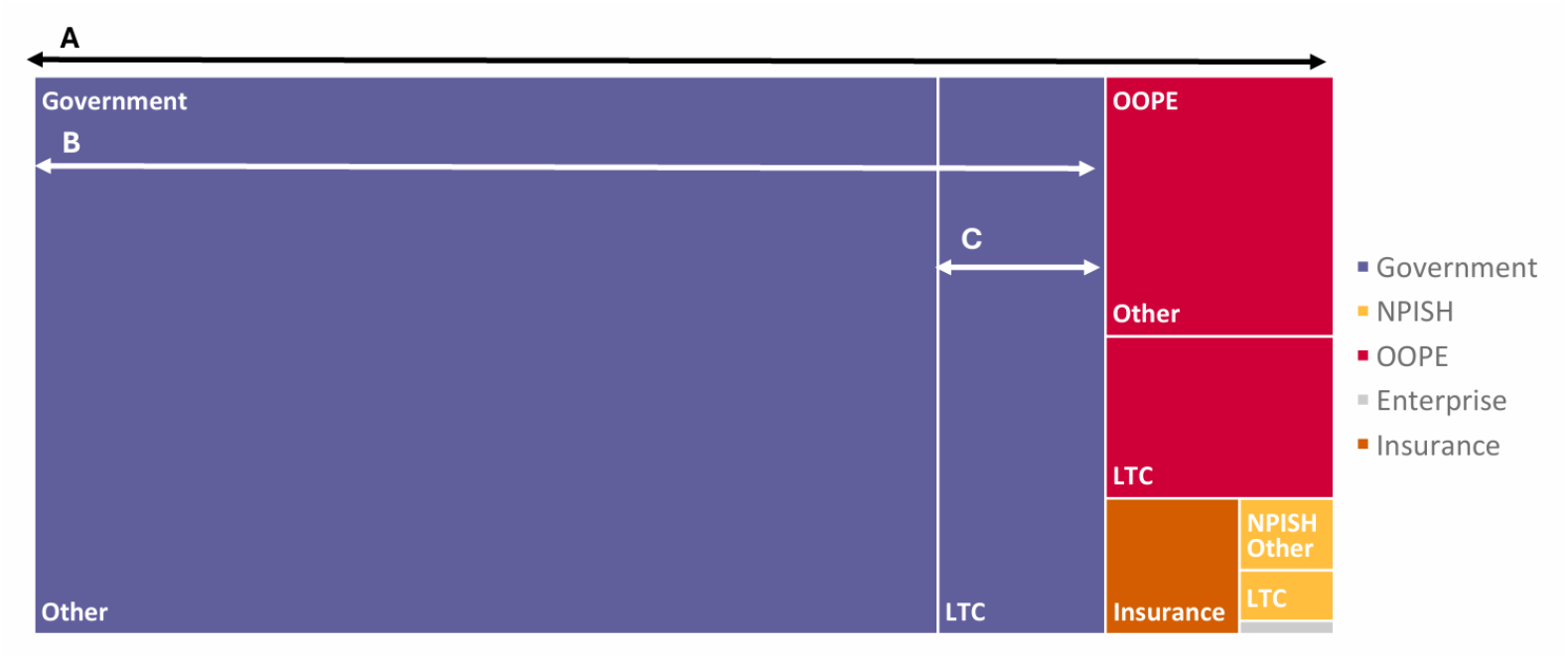
Treemap illustrating the composition of total UK healthcare expenditure. (A) total national healthcare expenditure (£296.6 billion); (B) total government-financed healthcare expenditure (£244.5 billion); (C) government-financed health-related long-term care expenditure (£34.3 billion). H-LTC=health-related long-term care; OOPE=out-of-pocket expenditure; NPISH=non-profit institutions serving households; Other=spending on healthcare functions excluding H-LTC. Source: UK Health Accounts.

The composition of this government-financed health-related long-term care expenditure is further detailed in *Figure 3*. Spending on inpatient care comprised almost two-thirds (£22.6 billion; 65.9%) of government-financed health-related long-term care expenditure, the majority of which (£21.2 billion; 61.8%) was provided by long-term care facilities such as nursing and residential homes *(Figure 3A)*. The remainder of spending on inpatient care was contributed by hospital inpatient services (£1.4 billion; 4.1%) *(Figure 3B)*. Home-based care accounted for approximately one-third of spending on health-related long-term care expenditure (£11.8 billion; 33.4%). This was largely spending by home care agencies (£11.3 billion; 32.9%) *(Figure 3C)*, with small amount of expenditure by community healthcare teams (£163.5 million; 0.5%) *(Figure 3D)*. Outpatient care was the smallest contributor to government-financed health-related long-term care expenditure, and included services provided by GPs (£211.5 million; 0.6%) *(Figure 3E)* and hospital outpatient departments (£48.7 million; 0.1%) *(Figure 3F*). Detailed annual figures for governmental spend on health-related long-term care by provider can be found in *Appendix 2 (supplemental material)*

**Figure 3:**
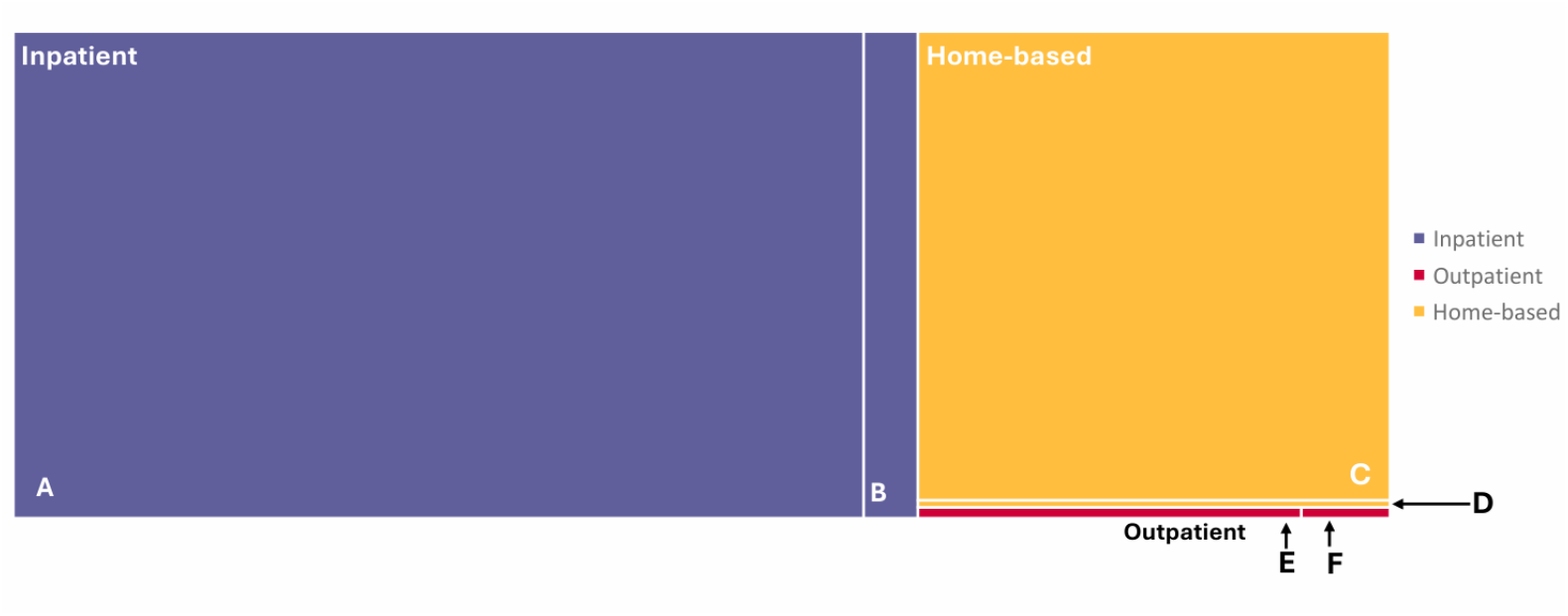
Treemap illustrating government-financed health-related long-term care expenditure in 2022 (in real terms, adjusted to 2023 prices). (A) long-term care facilities (£21.2 billion; 61.8%); (B) hospital inpatient services (£1.4 billion; 4.1%); (C) home care agencies (£11.3 billion; 32.9%); (D) NHS community health teams (£163.5 million;0.5%); (E) GPs (£211.5 million; 0.6%); (F) hospital outpatient services (£48.7 million; 0.1%). H-LTC=health-related long-term care; OOPE=out-of-pocket expenditure; NPISH=non-profit institutions serving households; Other=spending on healthcare functions excluding H-LTC. Source: UK Health Accounts.

#### (2) Trends in specialist palliative care expenditure

Specialist palliative care expenditure reported by Hospice UK grew from £1,027 million in 2013 to £1,137 million in 2022. This real terms growth of £110 million represents a proportional increase of 10.7% in specialist palliative care expenditure by Hospice UK member organisations over the ten-year period analysed. Real terms spending on specialist palliative care decreased in 2018, and in the years 2020 to 2022 consecutively. Detailed annual figures are available in *Appendix 3 (supplemental material)*.

## DISCUSSION

We demonstrate that real terms spending on government-financed health-related long-term care in the UK grew by 22.9% between 2013 and 2022, while real terms spending on specialist palliative care services in the UK grew by only 10.7%. It is not possible to identify direct spending on specialist and non-specialist palliative care within the UK Health Accounts data. We turned to Hospice UK accounts data in order to at least contextualise specialist palliative care spend; by any standard, this is a small figure in relation to overall long-term healthcare expenditure.

The majority of the UK’s specialist palliative care is provided by hospice charities. These organisations receive around one-third of their funding from government, generating their remaining income through fundraising activities. [17] This mixed funding model reflects those seen internationally, including in Germany, Hungary, Ireland, Malaysia, New Zealand, the Netherlands, Poland, Romania, Spain, the USA, and Zimbabwe. [23, 24] Rising costs, unmatched by increases in statutory funding for palliative care services, have resulted in financial hardship and reduced service provision. [17]

Palliative care services face rising demand associated with population ageing and the increasing prevalence of complex long-term conditions. While half of people who die in the UK have contact with specialist palliative care teams within the last three months of life, it is estimated that 100,000 people die each year needing palliative care but not receiving it. [25] Additional resources are required to keep pace with demand, yet instead, funding has reduced in real terms over time.

Previous studies suggest specialist palliative care is cost effective when compared with standard care for individuals at the end of life. [24, 26, 27] Considering the proportionally small figure spent on specialist palliative care as compared to national government expenditure on health-related long-term care as a whole, a small increase in statutory funding could have a large impact on the delivery palliative care services in the UK. [13]

When considering funding of palliative care in a global context, it becomes clear that palliative care is a public health issue. It is estimated that over 56 million people need palliative care globally, of whom only 17.9% are located in Europe. [24] Disparities in the palliative care workforce internationally are also clear. The UK hospice sector employed 16,000 clinical staff in 2022-23, with 95,000 volunteers supporting delivery of care and support services. [28]

Meanwhile, an estimated 400,000 health care workers globally are involved in delivering palliative care with the support of 1.2 million volunteers. [24] The challenges to funding and delivery of palliative care in the UK, previously ranked as providing the best quality of palliative care in international comparisons, are only exacerbated in low-income countries, where the availability of palliative care is substantially more limited and dedicated funding for palliative care may not be available. [9, 24] While this paper can provide a framework for comparison of palliative care expenditure between high-income countries with similar funding models, these results may have limited applicability in countries where palliative care services are less developed.

Our analysis was limited by the low granularity of the UK Health Accounts dataset. We could not identify specific expenditure figures for health-related social care included in reported health-related long-term care figures. Ongoing developments in the linkage of health and social care datasets may allow future studies to address this issue. [11] Additionally, direct comparison between national spending on health-related long-term care and specialist palliative care services was limited due to differences in the scale of expenditure figures.

The use of the System of Health Accounts 2011, an internationally standardised reporting tool, allows international comparisons of macro-level UK Health Accounts data. However, a specific figure for expenditure on palliative care cannot be determined due to the nature and purpose of the dataset. Expenditure on specialist and non-specialist palliative care within the National Health Service, in hospital inpatient and outpatient services and community nursing services, could not be delineated from other expenditure within the category of health-related long-term care. It is therefore difficult to make international comparisons of palliative care expenditure with available data. This is reflective of challenges to health economics research internationally, as reported data is dependent on the funding models used. [23] Routine data collection of governmental and non-governmental spending on specialist and non-specialist palliative care services across the UK would provide decision makers with key information for planning and delivery of services.

## CONCLUSION

Despite increasing recognition of the importance of palliative care, chronic lack of funding has left specialist palliative care services struggling to meet rising demands. In this study, low granularity of UK national health expenditure accounts limits national and international comparisons of spending on palliative care. Improvements to national health expenditure reporting, including the linking of health and social care datasets, would benefit the development of policy and planning of service delivery related to specialist and non-specialist palliative care. However, it is clear that UK expenditure on specialist palliative care services has not kept pace with growth in UK expenditure on health-related long-term care. Increased investment is required if palliative care services are to adequately address population need.

## Data Availability

All data used in this analysis are publicly available. UK Health Accounts data are available via the Office for National Statistics website at https://www.ons.gov.uk/peoplepopulationandcommunity/healthandsocialcare/healthcaresystem/datasets/healthaccountsreferencetables. Hospice UK data are available from annual Hospice Accounts reports, published online at https://www.hospiceuk.org/innovation-hub/support-for-your-role/non-clinical-resources/finance-support-for-hospices/financial-benchmarking.

## STATEMENTS

### Data Availability Statement

All data used in this analysis are publicly available. UK Health Accounts data are available via the Office for National Statistics website. [16] Hospice UK data are available from annual Hospice Accounts reports, published online. [17]

### Ethics approval

Ethics approval was not required. No human participants or patient identifiable data were involved.

### Patient consent for publication

Not applicable.

## Acknowledgements

We are grateful to Craig Duncan, Chief Operating Officer at Hospice UK for providing data on hospice expenditure.

We also thank the UK Health Accounts Team for providing additional information on the inclusion of hospice and specialist palliative care expenditure in the UK Health Accounts.

This research was supported by the Wolfson Palliative Care Research Centre, Hull York Medical School, University of Hull.

This work was undertaken as part of a UK Foundation Programme Office (UKFPO) Specialised Foundation Programme research placement.

## Contributors

FM supervised the project and was responsible for conceptualisation. ED analysed the data and wrote the first draft of the manuscript. All co-authors contributed to the interpretation of the results, and the drafting and approval of the final manuscript. FM is guarantor, accepts full responsibility for the finished work and/or the conduct of the study, and controlled the decision to publish.

## Funding

FM is a National Institute for Health and Care Research (NIHR) Senior Investigator.

ED is a NIHR Academic Clinical Fellow in Palliative Medicine.

The views expressed in this article are those of the author(s) and not necessarily those of the NIHR or the Department of Health and Social Care.

## REFERENCES

1. Sleeman KE, de Brito M, Etkind S, et al. The escalating global burden of serious healthrelated suffering: projections to 2060 by world regions, age groups, and health conditions. The Lancet Global Health. 2019;7(7):e883–e92. doi:10.1016/S2214-109X(19)30172-X

2. Etkind SN, Bone AE, Gomes B, et al. How many people will need palliative care in 2040? Past trends, future projections and implications for services. BMC Medicine. 2017;15(1):102. doi: 10.1186/s12916-017-0860-2

3. Office for Budget Responsibility. Drivers of rising health spending [online]. 2015. https://obr.uk/box/drivers-of-rising-health-spending (accessed 29 March 2025)

4. Stoye G, Warner M, Zaranko B. The past and future of UK health spending. London: Institute for Fiscal Studies [online]. 2024. https://ifs.org.uk/publications/past-and-future-uk-health-spending (accessed 16 February 2025).

5. Jayawardana S, Cylus J, Mossialos E. It’s not ageing, stupid: why population ageing won’t bankrupt health systems. European Heart Journal - Quality of Care and Clinical Outcomes. 2019;5(3):195–201. doi: 10.1093/ehjqcco/qcz022

6. Office for National Statistics. Profile of the older population living in England and Wales in 2021 and changes since 2011 [online]. 2023. https://www.ons.gov.uk/peoplepopulationandcommunity/birthsdeathsandmarriages/ageing/articles/profileoftheolderpopulationlivinginenglandandwalesin2021andchangessince2011/2023-04-03 (accessed 30 March 2025).

7. Office for National Statistics. National population projections: 2018-based [online]. 2019. https://www.ons.gov.uk/peoplepopulationandcommunity/populationandmigration/populationprojections/bulletins/nationalpopulationprojections/2018based (accessed 30 March 2025).

8. Kingston A, Robinson L, Booth H, et al. Projections of multi-morbidity in the older population in England to 2035: estimates from the Population Ageing and Care Simulation (PACSim) model. Age Ageing. 2018;47(3):374–80. doi: 10.1093/ageing/afx201

9. The Economist Intelligence Unit. The 2015 Quality of Death Index [online]. 2015. https://impact.economist.com/health/2015-quality-death-index (accessed 30 March 2025).

10. Hughes J. UK: the best place in the world to die. BMJ. 2015:h5440. doi: 10.1136/bmj.h5440

11. Cummins L, Julian S, Georghiou T, et al. Public expenditure in the last year of life. Nuffield Trust [online]. 2025. https://www.nuffieldtrust.org.uk/research/public-expenditure-in-the-last-year-of-life (accessed 29 March 2025).

12. Marie Curie. How many people need palliative care? Updated estimates of palliative care need across the UK, 2017-2021 [online]. 2023. https://www.mariecurie.org.uk/globalassets/media/documents/policy/policy-publications/2023/how-many-people-need-palliative-care.pdf (accessed 29 March 2025).

13. May P, Nikram E, Johansson T, et al. Specialist palliative care improves patient experience, reduces bed days and saves money: an economic modelling study of home- and hospital-based care. MedRxiv [Preprint]. 2025:2025.08.19.25333960. doi: 10.1101/2025.08.19.25333960

14. Office for National Statistics. UK Health Accounts: methodological guidance [online]. 2023. https://www.ons.gov.uk/peoplepopulationandcommunity/healthandsocialcare/healthcaresystem/methodologies/ukhealthaccountsmethodologicalguidance (accessed 15 January 2025).

15. Newson N. Hospices: State Funding. In Focus. House of Lords Library [online]. 2024. https://lordslibrary.parliament.uk/hospices-state-funding/ (accessed 29 March 2025).

16. Office for National Statistics. UK Health Accounts dataset: 2023 edition [dataset]. 2024. https://www.ons.gov.uk/peoplepopulationandcommunity/healthandsocialcare/healthcaresystem/datasets/healthaccountsreferencetables.

17. Hospice UK. Financial benchmarking: Hospice Accounts Reports [online]. 2025. https://www.hospiceuk.org/innovation-hub/support-for-your-role/non-clinical-resources/finance-support-for-hospices/financial-benchmarking (accessed 30 March 2025).

18. Office for National Statistics. Introduction to health accounts [online]. 2016. https://www.ons.gov.uk/peoplepopulationandcommunity/healthandsocialcare/healthcaresystem/methodologies/ (accessed 15 January 2025).

19. Office for National Statistics. Gross Domestic Product (GDP) [online]. 2025. https://www.ons.gov.uk/economy/grossdomesticproductgdp (accessed 06 January 2026).

20. OECD, Eurostat, WHO. A System of Health Accounts 2011: Revised edition. 2017. doi: 10.1787/9789264270985-en

21. Pask S, Murtagh FEM, Boland JW. Palliative care: what’s the evidence? Clinical Medicine. 2025;25(4):100320. doi: 10.1016/j.clinme.2025.100320

22. Office for National Statistics. CPI INDEX 06 : HEALTH 2015=100 [dataset]. 2025. https://www.ons.gov.uk/economy/inflationandpriceindices/timeseries/d7bz/mm23

23. Groeneveld EI, Cassel JB, Bausewein C, et al. Funding models in palliative care: Lessons from international experience. Palliative Medicine. 2017;31(4):296–305. doi: 10.1177/0269216316689015

24. Connor S. Global Atlas of Palliative Care 2nd Edition [online]. 2020. https://www.paho.org/en/node/75063 (accessed 12 September 2025).

25. Marie Curie. Better End of Life 2024. Time to care: Findings from a nationally representative survey of experiences at the end of life in England and Wales [online]. 2024. https://www.mariecurie.org.uk/document/experiences-at-the-end-of-life-in-england-and-wales (accessed 30 March 2025).

26. Clarke G, May P, Cook A, et al. Costs and cost-effectiveness of adult palliative and end-of-life care: Evidence briefing summary. London: National Institute for Health and Care Research (NIHR) Policy Research Unit (PRU) for Palliative and End-of-Life Care [online]. 2025. https://www.kcl.ac.uk/nmpc/assets/research/costs-and-cost-effectiveness-of-adult-palliative-and-end-of-life-care-evidence-briefing-summary.pdf (accessed 16 January 2026).

27. Naoum P, Athanasakis K, Pavi E. Is Palliative Care Cost-Effective? A Systematic Review of the Literature. Home Health Care Management & Practice. 2024;36(3):205–16. doi: 10.1177/10848223231209316

28. Hospice UK. Key facts about hospice care [online]. 2025. https://www.hospiceuk.org/about-us/key-facts-about-hospice-care (accessed 26 March 2025).

