## Supplemental Material for "How does palliative care fit into national health spending? A secondary analysis of trends in long-term healthcare expenditure in the United Kingdom"

#### Appendix 1: Structure of UK Health Accounts and Inclusion Decisions

Expenditure data is organised by three categories in the UK Health Accounts – financing scheme, provider organisation and healthcare function. [1-3] This appendix expands the categorisation of healthcare expenditure found in these accounts and explains in more detail the inclusion decisions for variables used in our analysis.

**Healthcare function:**

The UK Health Accounts include seven categories of healthcare function:

- Curative care
- Rehabilitative care
- Health-related long-term care (also referred to as Long-Term Care (Health))
- Ancillary services (including ambulance and patient transport)
- Medical goods (including pharmaceuticals, durable and non-durable goods and therapeutic appliances)
- Preventive care
- Governance, health system financing and administration

We selected health-related long-term care as our variable of interest given our objectives of measuring trends in health-related long-term care, and because this variable includes (but is not limited to) expenditure related to palliative care services.

**Financing scheme**:

The UK Health Accounts dataset categorises expenditure according to funding source, also termed financing scheme:

- Government
- Non-profit institutions serving households (charities)
- Voluntary insurance schemes (voluntary insurance claims from private medical insurance and employer self-insurance schemes)
- Enterprise (provision or purchase of healthcare services by companies for employees)
- Out-of-pocket payments (direct consumer spending on healthcare goods and services, such as prescription charges and direct payments for private healthcare services).

We considered only government-financed (public) spending on health-related long-term care in our analysis. This includes (but is not limited to) spending on NHS-commissioned specialist palliative care services. It excludes government grants to hospice charities and charitably funded specialist palliative care provided in hospices.

**Provider organisation:**

Expenditure is categorised by provider organisation in the UK Health Accounts:

- Hospitals
- Residential long-term care facilities (nursing homes, residential homes and hospices)
- Providers of ambulatory care (offices of general medical practitioners, dental practices, providers of home healthcare services, and other ambulatory care providers such as community health services)
- Ambulatory care providers (ambulance services)
- Retailers of medical goods
- Providers of preventive care
- Providers of healthcare system administration and financing
- Rest of the economy (households receiving Carer's Allowance benefits, and secondary providers of healthcare such as schools and policing services)

We identified providers of health-related long-term care services who provide services including (bit not limited to) palliative care using UK Health Accounts methodological guidance, with further information provided by the UK Health Accounts Team. We included hospitals and long-term care facilities providing inpatient care, offices of general medical practitioners and hospitals providing outpatient care, and home healthcare agencies and community nursing teams providing home-based care in our analysis (see *Supplementary Table 1*).

**Modes of provision:**

In the UK Health Accounts, health-related long-term care can be further categorised by the type of service provided – whether it is inpatient, outpatient, day or home-based care.

- **Day care:** refers to planned formal admissions to hospitals with the intention to discharge patients home the same day. As palliative care services *in hospitals* tend to operate outpatient clinic services rather than day case services in secondary care, this data was excluded from our analysis. Palliative day services in hospices are not explicitly included in this dataset.
- **Inpatient care:** refers to formal admissions to care facilities expected to involve an overnight stay. Includes care provided by hospitals and long-term care facilities (residential and nursing homes, and NHS-commissioned hospice services). This included local authority spending on packages of care which primarily provided support with basic activities of daily living required due to a health condition. It also included NHS Continuing Healthcare funding, which is NHS-funded social care required due to a primary health need.
- **Home-based care:** refers to care provided in a patient's home with a healthcare provider present. This includes NHS and local-authority funded home healthcare providers, such as home care agencies. Similar to long-term care facilities, this refers to spending on packages of care which primarily provide support with basic activities of daily livings required due to an underlying health condition, and NHS Continuing Healthcare funding. It also includes home-based care provided by 'other ambulatory providers', which refers to NHS community health services such as district nursing and community specialist palliative care nursing teams. We excluded government expenditure on Carer's Allowance, a means-tested benefit provided to informal carers, as this data was unlikely to include expenditure directly related to palliative care.

- **Outpatient care:** refers to care received on a healthcare providers' premises, where the patient is not formally admitted and is not expected to stay overnight. We included outpatient care provided in hospitals in our analysis, as some hospitals run palliative care outpatient clinics We also included care provided by GP surgeries, which deliver health-related long-term care services including (among other things) patient with life-limiting conditions.

***Supplemental Table 1:*** *Summary of inclusion decisions. All expenditure listed here refers to government-funded health-related long-term care. Palliative care refers to both specialist and non-specialist palliative care services. Source: UK Health Accounts [2,4]*

| **Mode of provision** | | **Provider** | | **Inclusion Status** | **Rationale** |
|---|---|---|---|---|---|
| HC31 | Long-term inpatient care | HP1 | Hospitals | Included | Includes specialist and non-specialist palliative care provided in hospitals. |
| | | HP2 | Residential long-term care facilities | Included | Includes NHS- and local authority- funded care in residential and nursing homes. This includes NHS-commissioned hospice inpatient services. Also includes NHS Continuing Healthcare funding. |
| HC32 | Long-term day care | HP1 | Hospitals | Excluded | Healthcare requiring formal admission but no overnight stay. Unlikely to include expenditure directly related to palliative care. |
| HC33 | Long-term outpatient care | HP1 | Hospitals | Included | Some hospital-based secondary palliative care teams provide outpatient clinics. |
| | | HP31 | Offices of general medical practitioners | Included | Includes spending on non-specialist palliative care provided by GPs, as well as for other conditions such as dementia and chronic obstructive pulmonary disease. |
| HC34 | Long-term home-based care | HP35 | Providers of home healthcare services | Included | Includes spending on home care services, such as those provided by home care agencies, funded by the local authorities and the NHS. An element of this will relate to spending on palliative care. Also includes NHS Continuing Healthcare funding. |
| | | HP3x | Other ambulatory providers | Included | Primarily consists of NHS-funded community services including district and specialist palliative care nursing teams. Includes some long-term disability services provided by the NHS. |
| | | HP81 | Households as providers of long-term care | Excluded | Includes government expenditure on Carer’s Allowance, a means tested benefit payment which supports informal carers. Excluded as there is no current guidance as to what proportion, if any, of this expenditure is related to palliative care services |

### Appendix 2: Analysis of long-term care expenditure in the UK – additional tables

***Supplemental Table 2:*** *Annual government spending on health-related long-term care services including (but not solely limited to) palliative care. All figures are presented in real terms adjusted to 2023 prices. Figures may not sum due to rounding. Source: UK Health Accounts.*

| Year | Inpatient care | | Outpatient care | | Home-based care | | Total |
|---|---|---|---|---|---|---|---|
| | Hospitals | Long-term care facilities | Hospitals | GP surgeries | Home care providers | NHS community services | |
| | £ billions | £ billions | £ millions | £ millions | £ billions | £ millions | £ billions |
| 2013 | 1.0 | 17.9 | 50.6 | 162.2 | 8.6 | 89.9 | 27.9 |
| 2014 | 1.1 | 17.9 | 51.4 | 180.3 | 9.1 | 93.8 | 28.4 |
| 2015 | 1.2 | 18.3 | 53.2 | 204.0 | 9.4 | 88.1 | 29.3 |
| 2016 | 1.3 | 18.6 | 53.7 | 168.1 | 9.6 | 93.4 | 29.9 |
| 2017 | 1.3 | 18.7 | 45.6 | 154.0 | 9.6 | 99.8 | 29.9 |
| 2018 | 1.2 | 19.3 | 41.3 | 157.4 | 9.8 | 102.0 | 30.7 |
| 2019 | 1.2 | 19.4 | 41.0 | 155.9 | 10.0 | 112.3 | 30.9 |
| 2020 | 1.3 | 21.4 | 44.8 | 249.0 | 11.0 | 118.0 | 34.1 |
| 2021 | 1.3 | 21.4 | 45.5 | 226.0 | 11.1 | 126.3 | 34.2 |
| 2022 | 1.4 | 21.2 | 48.7 | 211.5 | 11.3 | 163.5 | 34.3 |

### Appendix 3: Analysis of specialist palliative care expenditure - additional figures and tables

***Supplemental Table 3:*** *Annual hospice sector charitable expenditure in £ millions adjusted for inflation to 2023 prices. Source: Hospice UK.*

| Year | Hospice sector charitable expenditure £ millions |
|---|---|
| 2013 | 1,027.0 |
| 2014 | 1,067.3 |
| 2015 | 1,090.8 |
| 2016 | 1,113.1 |
| 2017 | 1,129.2 |
| 2018 | 1,125.2 |
| 2019 | 1,132.7 |
| 2020 | 1,148.8 |
| 2021 | 1,140.3 |
| 2022 | 1,137.0 |

### REFERENCES

1. Office for National Statistics. UK Health Accounts dataset: 2023 edition [dataset]. 2023. https://www.ons.gov.uk/peoplepopulationandcommunity/healthandsocialcare/healthcaresystem/datasets/healthaccountsreferencetables.
2. Office for National Statistics. UK Health Accounts: methodological guidance [online]. 2023. https://www.ons.gov.uk/peoplepopulationandcommunity/healthandsocialcare/healthcaresystem/methodologies/ukhealthaccountsmethodologicalguidance (accessed 15 January 2025).
3. OECD, Eurostat, WHO. A System of Health Accounts 2011: Revised edition. 2017. doi: 10.1787/9789264270985-en
4. Office for National Statistics. Introduction to health accounts [online]. 2016. https://www.ons.gov.uk/peoplepopulationandcommunity/healthandsocialcare/healthcaresystem/methodologies/ (accessed 15 January 2025).
